# On evaluation metrics for medical applications of artificial intelligence

**DOI:** 10.1101/2021.04.07.21254975

**Authors:** Steven A. Hicks, Inga Strümke, Vajira Thambawita, Malek Hammou, Michael A. Riegler, Pål Halvorsen, Sravanthi Parasa

**Affiliations:** SimulaMet, Oslo, Norway; Oslo Metropolitan University, Oslo, Norway; Swedish Medical Center, Seattle, USA

**Author notes:** contact/corresponding author.

**Keywords:** Gastroenterology, standardization, machine learning, evaluation metrics

## Abstract

Clinicians and model developers need to understand how proposed machine learning (ML) models could improve patient care. In fact, no single metric captures all the desirable properties of a model and several metrics are typically reported to summarize a model’s performance. Unfortunately, these measures are not easily understandable by many clinicians. Moreover, comparison of models across studies in an objective manner is challenging, and no tool exists to compare models using the same performance metrics. This paper looks at previous ML studies done in gastroenterology, provides an explanation of what different metrics mean in the context of the presented studies, and gives a thorough explanation of how different metrics should be interpreted. We also release an open source web-based tool that may be used to aid in calculating the most relevant metrics presented in this paper so that other researchers and clinicians may easily incorporate them into their research.

---

Improving healthcare applications and supporting decision making for medical professionals using methods from Artificial Intelligence (AI), specifically Machine Learning (ML), is a rapidly developing field with numerous retrospective studies being published every week. We also observe an increasing number of prospective studies involving large multi-center clinical trials testing ML systems’ suitability for clinical use. The technical and methodological maturity of the different areas varies, radiology and dermatology being examples of the more advanced ones^1^. In addition to these two examples, we observe a recent surge of studies in the field of gastroenterology^2^. Therefore, the present study focuses on the current development in gastroenterology due to its timeliness. However, our discussions, recommendations, and proposed tool are valid and useful in every clinical field adopting and employing ML-based systems.

The use of ML in gastroenterology is expected to significantly improve detection and characterization of colon polyps and other precancerous lesions of the Gastrointestinal (GI) tract^3^. These potential advances are mainly expected from artificial neural networks, specifically deep learning-based methods^4^. Safe and efficient adoption of ML tools in clinical gastroenterology requires a thorough understanding of the performance metrics of the resulting models and confirmation of their clinical utility^5^.

Creating strong evidence for the usefulness of ML models in clinical settings is an involved process. In addition to the relevant epidemiological principles, it requires a thorough understanding of the properties of the model itself and its performance. However, despite increased interest in ML as a medical tool, understanding of how such models work and how to aptly evaluate them using different metrics is widely lacking. In this article, we use examples of metrics and evaluations drawn from a variety of peer-reviewed and published studies in gastroenterology to provide a guide explaining different evaluation metrics, including how to interpret them. Note that we do not discuss the quality of these studies, but merely use them to discuss how different metrics give different interpretations of the quality of an ML model.

The main contributions are: We present a detailed discussion on metrics commonly used for evaluating ML classifiers, examine existing research using ML in gastroenterology along with reported metrics, and we discuss the different metrics’ interpretations, usefulness, and shortcomings. To this end, we recalculate the reported metrics and calculate additional ones to further analyze the performance of the presented methods. Additionally, we present a web-based open source tool intended to let researchers perform metrics calculations easily, both for their own and other reported results to allow for comparison. The tool is accessible via www.medimetrics.no, and the source code via github.com/simula/medimetrics.

## Metrics

The relevant quantities for calculating the metrics for a binary classifier are the four entries in the confusion matrix

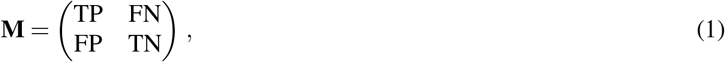

which are introduced below. Note that we limit ourselves to binary classification in this study, which is at the time of writing most common for medical applications, e.g., cancer/no-cancer or polyp/no-polyp. Some metrics have different interpretations in the context of evaluating multi-class classification methods. Although these discussions fall outside the scope of the present one, the underlying principles still apply in multi-class settings. Furthermore, some methods have their metrics calculated on a per-finding basis, meaning there can be multiple instances for one image, and hence more positive samples than total samples (e.g., images or videos) in a data set.

### True Positive (TP)

The true positive denotes the number of correctly classified positive samples. For example, the number of frames containing a polyp correctly predicted as having a polyp.

### True Negative (TN)

The true negative denotes the number of correctly classified negative samples. For example, the number of frames not containing a polyp correctly predicted as not having a polyp.

### False Positive (FP)

The false positive denotes the number of samples incorrectly classified as positive. For example, the number of frames not containing a polyp incorrectly predicted as having a polyp.

### False Negative (FN)

The false negatives denotes the number of samples incorrectly classified as negative. For example, the number of frames containing a polyp incorrectly predicted as not having a polyp.

### Accuracy (ACC)

The accuracy is the ratio between the correctly classified samples and the total number of samples in the evaluation data set. This metric is among the most commonly used in applications of ML in medicine, but is also known for being misleading in case of different class proportions, since simply assigning all samples to the prevalent class is an easy way of achieving high accuracy. The accuracy is bounded to [0, 1], where 1 represents predicting all positive and negative samples correctly, and 0 represents predicting none of the positive or negative samples correctly.

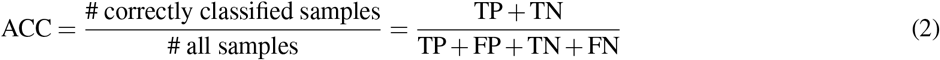

### Recall (REC)

The recall, also known as the sensitivity or True Positive Rate (TPR), denotes the rate of positive samples correctly classified, and is calculated as the ratio between correctly classified positive samples and all samples assigned to the positive class. The recall is bounded to [0, 1], where 1 represents perfectly predicting the positive class, and 0 represents incorrect prediction of all positive class samples. This metric is also regarded as being among the most important for medical studies, since it is desired to miss as few positive instances as possible, which translates to a high recall.

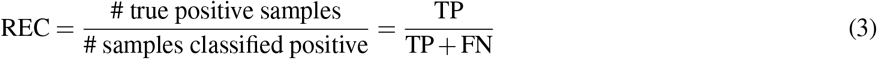

### Specificity (SPEC)

The specificity is the negative class version of the recall (sensitivity) and denotes the rate of negative samples correctly classified. It is calculated as the ratio between correctly classified negative samples and all samples classified as negative. The specificity is bounded to [0, 1], where 1 represents perfectly predicting the negative class, and 0 represents incorrect prediction of all negative class samples.

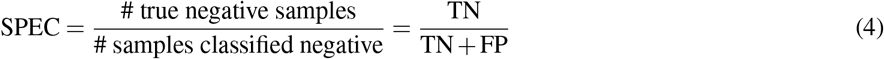

### Precision (PREC)

The precision denotes the proportion of the retrieved samples which are relevant and is calculated as the ratio between correctly classified samples and all samples assigned to that class. The precision is bounded to [0, 1], where 1 represents all samples in the class correctly predicted, and 0 represents no correct predictions in the class.

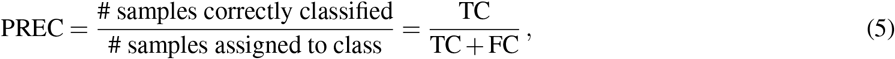

where *C* denotes “class”, and can in binary classification be either positive (*P*) or negative (*N*). The terms precision and Positive Predictive Value (PPV) are often used interchangeably.

### F1 score (F1)

The F1 score is the harmonic mean of precision and recall, meaning that it penalizes extreme values of either. This metric is not symmetric between the classes, i.e., it depends on which class is defined as positive and negative. For example, in the case of a large positive class and a classifier biased towards this majority, the F1 score, being proportional to TP, would be high. Redefining the class labels so that the negative class is the majority and the classifier is biased towards the negative class would result in a low F1 score, although neither the data nor the relative class distribution have changed. The F1-score is bounded to [0, 1], where 1 represents maximum precision and recall values and 0 represents zero precision and/or recall.

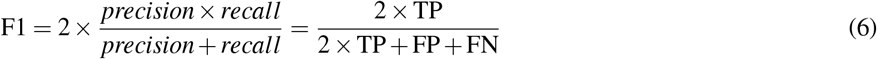

### Matthews Correlation Coefficient (MCC)

Pearson’s correlation coefficient^6^, takes on a particularly simple form in the binary case. This special case has been coined the MCC^7^, and become popular in ML settings for its favorable properties in cases of imbalanced classes^8^. It is essentially a correlation coefficient between the true and predicted classes, and achieves a high value only if the classifier obtains good results in all the entries of the confusion matrix Equation 1. The MCC is bounded to [−1, 1], where a value of 1 represents perfect prediction, 0 random guessing and −1 total disagreement between prediction and observation.

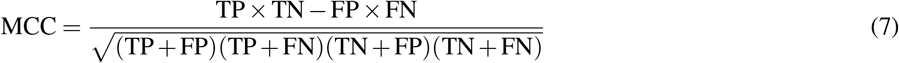

### PPV

The PPV is the ratio between correctly classified positive samples and all samples classified as positive, and equals the precision for the positive class. The PPV is bounded to [0, 1], where 1 represents all positive samples predicted correctly, and 0 represents no correct positive class predictions.

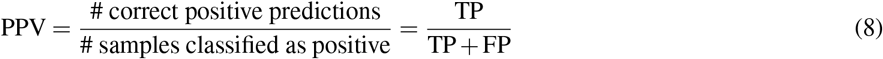

### Negative Predictive Value (NPV)

The NPV is the ratio between correctly classified negative samples and all samples classified as negative, and equals the precision for the negative class. The NPV is bounded to [0, 1], where 1 represents all negative samples predicted correctly, and 0 represents no correct negative class predictions.

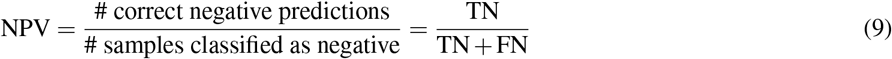

### Threat Score (TS)

The TS, also called the Critical Success Index (CSI), is the ratio between the number of correctly predicted positive samples against the sum of correctly predicted positive samples and all incorrect predictions. It takes into account both false alarms and missed events in a balanced way, and excludes only the correctly predicted negative samples. As such, this metric is well suited for detecting rare events, where the model evaluation should be sensitive to correct classification of rare positive events, and not overwhelmed by many correct identifications of negative class instances. The TS is bounded to [0, 1], where 1 represents no false predictions in either class, and 0 represents no correctly classified positive samples.

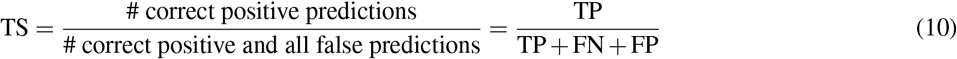

We do not consider the AUROC (Area under the Receiver Operating Characteristic Curve) or AUPRC (Area under the Precision Recall Curve) since these cannot be calculated without access to the model, or from the entries of the confusion matrix. Extensive research has been done on their usefulness, and we refer the interested reader to^9^.

#### Class mixture

Binary classification problems can be expressed in terms of a mixture model, the total data distribution modelled as

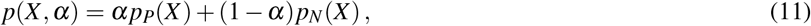

where *X* represents data samples, *p*_*P/N*_ denotes the positive/negative class distributions, and *α* the mixture parameter of the positive class, calculated as 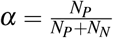, with *N*_*P/N*_ the total number of positive/negative class data samples. Studies in which the classification threshold of model outputs are tuned using a class imbalanced data set, should investigate how these perform on other class admixtures. This is an important step to assess whether bias towards either class has been introduced and to what extent.

#### Blinded data

Model development is typically split into three phases. First, the model is trained on a training dataset appropriate for the given task. Second, during training, the model is continuously validated on data not part of the training data, to evaluate the model’s performance on unseen data. Last, after the model has finished training, it is tested on a test dataset for which the final metrics should be calculated. Regardless of which metric is used, this can only be as informative as the classifier’s performance on the test data. Blinding data, i.e., withholding data from those performing the experiment, is an important tool in many research fields, such as medicine. In some experiment types, it is difficult to achieve blinding, but the analysis in a setting where the data has already been collected can almost always be blinded. Misconceptions regarding the objectivity of statistical analysis should not keep researchers from blinding the data^10^. For ML analyses, such as the ones described in the present work, this means that one should set aside representative data that can be used for testing after the training and tuning processes are finished.

## Methods

In the following, we identify a subset of relevant studies for our analysis. Medical studies presenting ML applications often refer to them simply as “AI systems”. While AI has certainly received an unprecedented amount of attention over the past years, and presenting systems using this term emphasizes their novelty, the term is imprecise. Hence, we refrain from using this generic term in the following and instead refer to the exact model architecture used.

### Study selection

The studies used for this work are chosen based on the following rational considerations. Our starting point is a recent review of AI in gastroenterology^11^. The review contains 138 articles, from which we select five studies that represent existing work using ML in gastroenterology. The selection criteria are as follows

i. Report sufficient information (many studies report so few metrics that it is not possible to calculate other metrics) for reproducing the reported metrics and calculating metrics not reported.
ii. Represent different cases of interest for performance metrics discussions.

In addition, we select a recent study reporting results from a large clinical trial, which was not included in the aforementioned review. The following contains a brief description of the selected studies and reported metrics.

#### Study 1

Hassan et al.^12^ introduce an unspecified “AI system” called GI-Genius to detect polyps, trained and validated using 2, 684 videos from 840 patients collected from white-light endoscopy. All 840 patients are randomly split into two separate data sets, one for training and one for validation. The validation data set contains 338 polyps from 105 patients, where 168 of the identified polyps are either adenomas or sessile serrated adenomas. The authors report a sensitivity of 99.7% as the main performance metric, which is calculated from 337 TPs out of the total 338 positive samples. From this, readers are likely to conclude that only one FN instance is identified in the validation set. No other metrics are reported, and while it is reported that each colonoscopy contains 50, 000 frames, no further details are given on the exact number of frames per video.

#### Study 2

Mossotto et al.^13^ use several ML models to classify diseases commonly found in the GI tract, using endoscopic and histological data. The data consist of 287 patients, from which 178 cases are Crohn’s Disease (CD), 80 cases are Ulcerative Colitis (UC), and 29 cases are Unclassified Inflammatory Bowel Disease (IBDU). Results are shown from unsupervised (clustering) and supervised learning. The latter is used to classify CD and UC patients. For this, the data is divided into a model construction set consisting of 210 patients (CD = 143, UC = 67), a model validation set of 48 patients (CD = 35, UC = 13), and an IBDU reclassification set containing 29 IBDU patients. The model is thus not trained on IBDU data, and the latter data set is excluded from the present discussion. The model construction set is stratified into a discovery set used to tune the parameters for CD versus UC discovery, and one for training and testing. For the best performing supervised model, tested on the test set, an accuracy of 82.7%, a precision of 0.91, a recall of 0.83 and an F1 score of 0.87 are reported, see Table 2 in^13^. On the validation set, the reported numbers are an accuracy of 83.3%, a precision of 0.86, a recall of 0.83, and an F1 score of 0.84, see Table 3 in^13^. These reported results are also listed in Table 1 under study 2.

**Table 1.** The reported metrics of the selected studies. The *STUDY* column represents each of the five studies selected for metric recalculation. The *SET* column is the different metrics calculated for the same set of data. The *REPORTED* column is how the metrics were reported in the respective study. To refer to the tables in each respective paper, we use T to refer to the table number and R for the row number. The *EVALUATION* column is the method used to generate the metrics. The *TOTAL* column is the total number of samples used in the metrics calculations. The *POS* and *NEG* columns represent the total number of positive and negative samples, respectively. The remaining columns correspond the aforementioned metric acronyms described in the main text.

| STUDY | SET | REPORTED | EVALUATION | TOTAL | POS | NEG | TP | TN | FP | FN | ACC | PREC | REC | F1 | SPEC | MCC | NPV | TS |
| --- | --- | --- | --- | --- | --- | --- | --- | --- | --- | --- | --- | --- | --- | --- | --- | --- | --- | --- |
| 1 | 1 | In-text | Per-finding | - | 338 | - | 337 | - | - | 1 | - | - | 1.00 | - | - | - | - | - |
| 2 | 1 | T2.R1 | Per-frame | 210 | 143 | 67 | - | - | - | - | 0.71 | 0.89 | 0.68 | 0.75 | - | - | - | - |
|  | 1 | T2.R2 | Per-frame | 210 | 143 | 67 | - | - | - | - | 0.77 | 0.81 | 0.88 | 0.83 | - | - | - | - |
|  | 1 | T2.R3 | Per-frame | 210 | 143 | 67 | - | - | - | - | 0.87 | 0.91 | 0.84 | 0.87 | - | - | - | - |
|  | 2 | T3.R1 | Per-frame | 48 | 13 | 35 | - | - | - | - | - | 0.65 | 0.85 | 0.73 | - | - | - | - |
|  | 2 | T3.R2 | Per-frame | 48 | 35 | 13 | - | - | - | - | - | 0.94 | 0.83 | 0.88 | - | - | - | - |
|  | 2 | T3.R3 | Per-frame | - | - | - | - | - | - | - | 0.83 | 0.86 | 0.83 | 0.84 | - | - | - | - |
| 3 | 1 | T1 | Per-frame | 106 | - | - | 65 | 33 | 7 | 1 | 0.94 | 0.90 | 0.98 | - | 0.83 | - | 0.97 | - |
| 4 | 1 | T2.R1 | Per-frame | 6,000 | 6,000 | 0 | 5,663 | 0 | 251 | 337 | - | - | 0.94 | - | - | - | - | - |
|  | 1 | T2.R2 | Per-frame | 1,414 | 1,414 | 0 | 1,296 | 0 | 41 | 118 | - | - | 0.92 | - | - | - | - | - |
|  | 1 | T2.R3 | Per-frame | 21,572 | 0 | 21,572 | 0 | 20,691 | 1,004 | 0 | - | NA | - | - | 0.96 | - | - | - |
|  | 2 | T2.R4 | Per-frame | - | - | - | 570 | 0 | 42 | 76 | - | 0.88 | - | - | - | - | - | - |
|  | 3 | In-text | Per-frame | 60,914 | - | - | - | - | - | - | - | - | 0.92 | - | - | - | - | - |
|  | 4 | In-text | Per-frame | 1,072,483 | 0 | 1,072,483 | 0 | - | - | 0 | - | - | - | - | 0.95 | - | - | - |
| 5 | 1 | T1 | Per-frame | - | - | - | 3723 | 4735 | 262 | 930 | 0.88 | - | 0.80 | - | 0.95 | - | - | - |

**Table 2.**
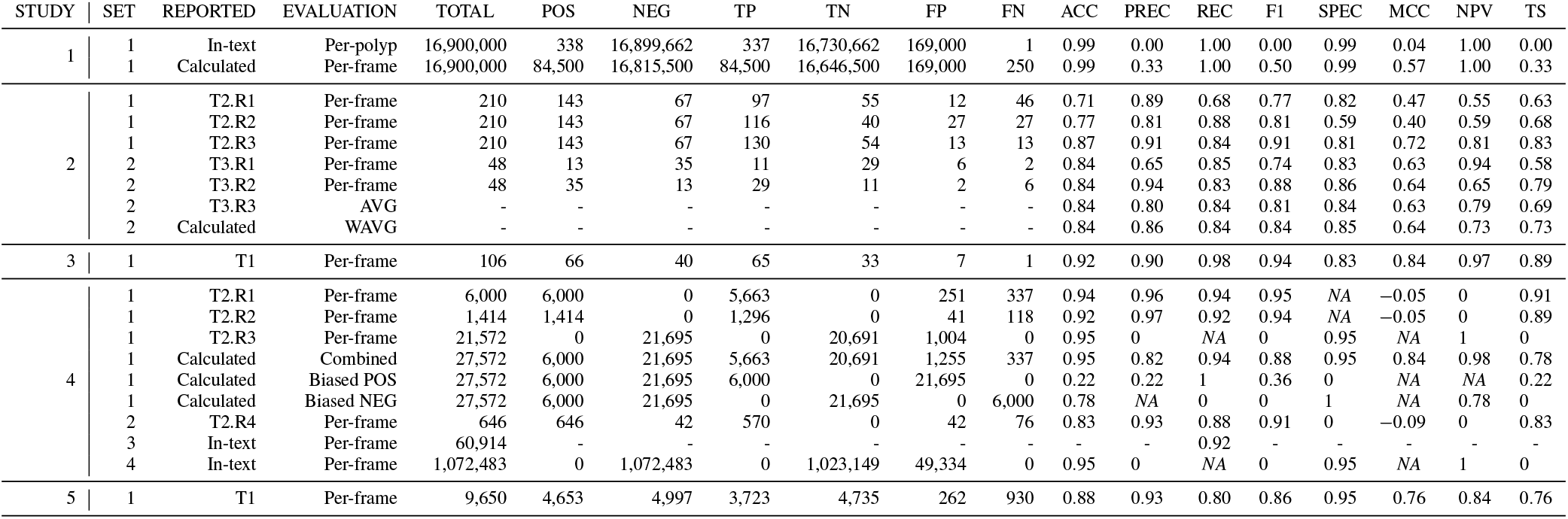
The recalculated metrics of the selected papers. Columns represent the same as described in Table 1.

#### Study 3

Byrne et al.^14^ introduce a Convolutional Neural Network (CNN) to differentiate diminutive adenomas from hyperplastic polyps. They define the four classes NICE type 1, NICE type 2, no polyp, and unsuitable. The training data set contains 223 polyp videos, consisting of 60, 089 frames in total, with 29% containing hyperplastic polyps, 53% containing adenomas polyps, and 18% containing no polyps. The model is tested on 158 videos, and 32 of these are removed due to the reported instances in the videos. Three are sessile serrated polyps, 25 are identified as normal tissue or lymphoid aggregate, two are fecal material, one video is corrupted, and two contain multiple polyp frames. The resulting 125 videos are used to evaluate the CNN model again, which is unable to confidently^1^ identify 19 of the 125 polyps. The 19 videos on which the model does not reach this confidence threshold are therefore removed from the test data set, and the model is evaluated using the remaining 106 videos. Finally, after this data filtering, the model achieves an accuracy of 94%, a sensitivity of 98%, a specificity of 83%, a PPV of 90% and an NPV of 97%, see Table 1 under study 3.

#### Study 4

Wang et al.^15^ present a near real-time deep learning-based system for detecting colon polyps using videos from colonoscopies. The model is trained on data collected from 1, 290 patients and validated on 27, 113 colonoscopy images from 1, 138 patients showing at least one detected polyp. It is then tested on a public database containing 612 images with polyps^16^. As the presented method is able to differentiate between different polyps within the same image, there may be more true positives than images in the data set. This is also the reason why the metrics are reported on a per-image basis. The reported results show that the method is highly effective, with a per-image-sensitivity of 94.38% and a per-image-specificity of 95.92%. As the metrics are reported separately for images containing polyps and those that do not, recalculating the metrics as presented provides an inaccurate representation of the model’s actual performance. This is because there are either no true positives or no true negatives, depending on the metrics used.

#### Study 5

Sakai et al.^17^ propose a CNN-based system to automatically detect gastric cancer in images from colonoscopies. The model is trained on a data set of 172, 555 images containing gastric cancer and 176, 388 images of normal colon. For evaluation, the model is tested on 4, 653 cancer images and 4, 997 normal images, on which it achieves an accuracy of 87.6%, a sensitivity of 80.0%, a specificity of 94.8% (see Table 1 in^17^), and a PPV of 93.4%. A method capable of distinguishing which regions of an image contain signs of gastric cancer is also presented. This method uses a sliding-window approach, where the model predicts the presence of gastric cancer in specific regions of the image to generate a block-like heat map covering the afflicted areas. This detection model is tested on 926 images, where it achieves an accuracy of 89.9% on cancer images and an accuracy of 70.3% on normal images.

## Results

In this section, we perform a recalculation of all reported and missing metrics in the selected studies. Based on this, we discuss the usefulness of different metrics and how to obtain a realistic and complete picture of the performance of a classifier. This is done by extracting reported numbers and metrics from each study and using these to calculate additional metrics, which gives additional perspectives on the possible evaluations and could lead to different conclusions. In some cases, assumptions must be made in order to calculate metrics or assess model performances under different conditions. All assumptions made in this study are detailed in the relevant discussions.

We also present our freely available online tool, which allows medical experts to calculate all presented metrics from classifier predictions, or those which can be calculated from a subset of metrics. This can be used for a variety of different usage scenarios, like gaining a better understanding of studies using ML classifiers, calculate missing metrics for studies which do not report them, to double-check calculations, and to calculate metrics for new studies.

### Precision and recall

To reproduce the results of *Study 1*^12^, it is necessary to make some assumptions. Primarily, no information is given regarding the total number of frames for all videos, but as an average of 50, 000 frames per video is reported, we use this to calculate the total number. Further, we calculate two sets of metrics, see Table 2 under study 1. For the first row, we calculate TP=337, TN=16, 730, 662, FP=169, 000, and FN=1 using the same per polyp detection evaluation as *Study 1*. In the second row, we assume that ten seconds around the polyp are either detected correctly or missed with a frame rate of 25 fps. This yields TP=84, 500, TN=16, 646, 500, FP=169, 000, and FN=250, which are used in our calculations. FP for both calculations are obtained based on the reported 1% FPs per video, i.e., 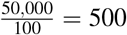. These assumptions yield two sets of results for the evaluation, which, if considered jointly, give a more thorough understanding of the performance. In any case, the reported values are not sufficient for reproducibility without making assumptions.

Assuming the most optimistic case amounts to mixture components of 0.995 and 0.005 for the positive and negative classes, respectively, meaning extremely imbalanced classes. The authors report a recall of 99.7%, in which case we calculate a precision of 0.33. Clearly, the recall must be interpreted with care in cases of strongly imbalanced classes. The reason is that precision and recall are both proportional to TP, but have an inverse mutual relationship: High precision requires low FP, so a classifier maximizing precision will return only very strong positive predictions, which can result in positive events missed. On the other hand, high recall is achieved by assigning more instances to the positive class, to achieve a low FN. Whether to maximize recall or precision depends on the application: Is it most important to identify *only* relevant instances, or to make sure that all relevant instances are identified? Regardless of which is the case, this should be clearly stated, and both metrics should be reported. The balance between the two has to be based on the medical use case and associated requirements. For example, some false alarms are acceptable in cancer detection, since it is crucial to identify all positive cases. On the other hand, for the identification of less severe disease with high prevalence, it can be important to achieve the highest possible precision. A low precision combined with a high recall implies that the classifier is prone to set of false alarms (FPs), which can result in an overwhelming manual workload and time wasted.

### Mixture parameter dependent tuning

In *Study 2*, Mossotto et al.^13^ split the model construction data set into two subsets of equal size and class distribution, with mixture components 0.68 and 0.32 for the two classes CD and UC. One of these subsets is used to tune parameters to maximize CD versus UC classification, meaning that the classification task is done with the underlying assumption that the class admixture will remain constant. This is trivially true for the training and test data, being the other of the two subsets, but not for the validation set, where the corresponding mixture components are 0.73 and 0.27. The authors do not mention the deviation from the tuned admixture, nor do they investigate systematically how much a given deviation affects the reported performance metrics. Without access to the resulting model, this cannot be investigated further in this article or by other interested readers. Consequently, there is no way of knowing the sensitivity the presented method has to the class admixture.

The study does not report any of the confusion matrix entries, thus not facilitating the process of reproducible results. However, we can derive the TN by multiplying the total number of positive samples with the recall, TP = REC × (TP + FN). The FP are then obtained via 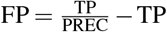. From this, we can calculate the reported and missing metrics. The authors report the positive class precision, which represents the PPV. In addition, the NPV should also have been reported for completeness. As shown in Table 2, the NPV is lower for the validation data sets where the precision is high, and vice versa. Calculating the MCC, this is stable around 0.63 over all validation sets listed in Table 3 of^13^, although not as high as any of the reported metrics.

### Blinded data

In *Study 3*, Byrne et al.^14^ remove data samples for which the classifier does not achieve high confidence, as well as videos with more than one polyp. They calculate the model’s performance for the different videos in the study, and remove the ones on which the model performs poorly. As such, the results reported from the study concerns a very specific selection of their data, made after the model has been adjusted. Excluding data on which a model performs poorly leads to a misrepresentation of its abilities and should not be done. If the classification task is too difficult or the removed data was faulty, this should instead be reported, and a new classifier should be trained for a more limited task. The metrics reported from a study should be calculated *after* the final model calibration and subsequent testing on blinded data.

### Negative and positive class performance

In *Study 4*, Wang et al.^15^ report high per-image-sensitivity (recall) values of 0.94, 0.92 and 0.92, see Table 2 in^15^, or metric set 1 under study 4 in Table 1. For the first two of these, sufficient numbers are reported to reproduce the reported metrics, as well as to calculate the corresponding positive class precisions, which are reported as 0.96 and 0.97, respectively. In the third case, the positive class precision cannot be calculated since the positive and negative samples are separated for the test. No explanation or reason is given regarding why the tests are performed only on the separated classes and not together, which would give a better overall impression of the performance. For the first data set, it is not clear how the numbers are calculated, as no test set is mentioned. This could mean that the reported sensitivity is calculated on the training data.

Rows five to seven in Table 2 show the results achieved when combining the negative and positive class samples. Since we do not know if the obtained model is biased towards the negative or positive class, we present three evaluations: In row five, we assume that the positive and negative results can simply be combined, which gives an overall MCC of 0.84 and NPV of 0.98, indicating good performance. In rows six and seven, we assume that the model is biased towards the positive or negative class, respectively. The resulting MCCs are both −0.05 and the NPVs both 0. This means that the classifier, which seemed to perform exceptionally well based on the reported numbers, is actually severely under-performing on the negative class. Besides these ambiguities, the results for the first three data sets indicate strong performance, but using the same numbers to calculate metrics more sensitive to bias, reveals severe under-performance (see Tables 1 and 2 for all reported and calculated numbers).

While detection and classification are in principle the same task for a fixed number of instances per class, the study in^15^ faces a challenge: The negative class is unbounded, i.e., the number of negative instances is undefined. The more sensitive the classifier is, the larger the negative class effectively becomes, as the classifier generates FP instances, and the negative class instances can be calculated as Neg = TN + FP. In general, evaluating without clearly defining boundaries for the classes is risky, as it can lead to an unclear impression of the model performance, in either the positive or negative direction. It is also nearly impossible for follow up studies to reproduce and compare results.

Without a well-defined number of true negatives in a video (or set of images), each of the frames not containing a polyp and each of the pixels not being part of a polyp are in principle true negatives. Optimally, the classes should instead be balanced, at best with a mixture parameter of 0.5. If this is not possible, the study should at least be based on well-motivated assumptions informed by real-world properties. For example, a standard colonoscopy contains on average *n* number of frames and *m* polyps found per examination. Most colonoscopies take less than an hour, so assuming a 24 hour time frame would be an unreasonable assumption within such boundaries.

Keeping the positive and negative classes separated in an evaluation can lead to misleading results and can make a model appear very different in terms of performance, depending on the presentation. The most important question that one should ask before performing the evaluation is: Which evaluation and metrics provide the most accurate representation regarding how the model will perform in the real world? This needs to be an overarching picture including both classes, and a set of diverse and well-suited metrics.

### Class dependent performance

*Study 5*^17^ contains a confusion matrix, enabling us to calculate most metrics for the reported results. As shown in Table 1, reported accuracy is 0.88, the specificity 0.95, the sensitivity 0.80, and the PPV 0.93. The PPV indicates the precision for the positive class and should be accompanied with the corresponding metric for the negative class, i.e., the NPV, which we calculate to be 0.84. This could indicate that the model is better at classifying positive than negative samples correctly, which would be surprising, given that the model is trained on slightly more negative than positive class images. However, not knowing the loss function or which measure the model was optimized for, we cannot investigate this further. What we do know is that a large number of FNs directly cause the low NPV: On the test data set, the model has a significantly higher number of FNs, meaning missed detection, than FPs, meaning a false alarm and in this case over-detection of cancer. The study specifically states reducing misdetection to be the primary motivation for using ML assisted diagnosis, and should thus have reported metrics providing a more comprehensive representation of the model’s performance in this regard. For instance, the MCC, which measures the correlation between the true and predicted classes, and is high only if the prediction is good on the positive *and* the negative class. By using the reported results, we calculate the MCC to be 0.76, which is still an acceptable performance, although not as high as the metrics reported by^17^. Which metric values are acceptable depends on non-technical aspects, e.g., the human performance baseline or requirements from hospitals or health authorities.

A potential weakness associated with the NPV is its dependence on TNs, which can overwhelm a classifier whose purpose is detecting rare events. In such cases, the TS, which does not take TNs into account, can be advantageous. From the values reported in Table 1 of^17^, the TS value is 0.76, again indicating that the model performs sub-optimally with respect to the objective, despite achieving high accuracy and precision values. In conclusion, the reported metrics show that the model, for the most part, performs well on the evaluation data set. When taking the recalculated metrics into account, we see that the model is more prone to misdetection than causing false alarms.

### MediMetrics

Together with this study, we release a web-based tool called *MediMetrics* for calculating the metrics introduced in Table, to make them easily accessible for medical doctors and ML researchers alike. From the provided input, the tool automatically calculates all possible metrics and generates useful visualizations and comparisons the user may freely use in their research. The tool is open-source, and the code available on GitHub^2^.

## Discussion

There are many available metrics that can be used to evaluate binary classification models. Using only a subset could give a false impression of a model’s actual performance, and in turn, yield unexpected results when deployed to a clinical setting. It is therefore important to use a combination of multiple metrics and interpret the performance holistically. Calculating multiple metrics does not require extra work in terms of study design or time, thus there is no apparent reason not to include a set of metrics, besides lack of space, obfuscating actual performance, or lack of knowledge regarding classifier evaluation. Besides interpreting the different metrics together, metrics for the separate classes should be calculated individually. Special care should be taken in cases of imbalanced classes, and the robustness of the classifier’s performance tested over a range of class admixtures. In general, a high score in any metric should be regarded with suspicion.

Training and evaluation sets should be strictly separated: Optimally, the data should be split into training, validation, and test data sets. The test data set should be separate from the other partitions to avoid introducing bias on the parameters set during the tuning phase. Furthermore, data regarding the same instance should not be shared across data splits. For example, frames of the same polyp from different angels should not shared across the training and test data sets. Once the model’s performance has been optimized on the training data, including tests on a validation set, it can be finally evaluated using the test set. This last step should thus not involve additional tuning, and the test data should not be made available to the analysis before results are fixated for publication. We argue strongly that this should be the standard for studies on the performance of ML classifiers used in medicine in the future. If possible, cross-dataset testing should be performed, meaning in this context that the training and test data are obtained from different hospitals or at least different patients.

In general, all studies involving classification should report the obtained TP, FP, TN, and FN values for validation and test data. In addition, the data along with either the source code, the final models or both should be made available. If this is not possible, other alternatives, like performing additional evaluation on public data sets, such as Kvasir^18^ or the Sun database^19^), should be considered. If such an alternative is chosen, it is important to check if the test data is outside the distribution of the training data^20^, and in that case, re-fit the model’s parameters. Although public data sets do not match the purpose of the study, evaluating the model on such data, by either re-training it or applying it directly on the data if similar to the initial data used for training, would allow others to compare methods and results.

## Data Availability

Not applicable

## Author contributions

Steven A. Hicks, Inga Strümke, Vajira Thambawita, Michael A. Riegler, Pål Halvorsen, Malek Hammou and Sravanthi Parasa conceived the experiment(s).

Steven A. Hicks, Inga Strümke, Vajira Thambawita and Michael A. Riegler conducted the experiment(s).

Steven A. Hicks, Inga Strümke, Vajira Thambawita, Michael A. Riegler and Pål Halvorsen analyzed the results. All authors reviewed the manuscript.

## Footnotes

1 For each image, the model gives a confidence value ranging from 0 to 1. If the confidence level is below 0.5, the model is not considered confident enough to keep the prediction.

2 github.com/simula/medimetrics

